# Plasma Glial Fibrillary Acidic Protein (GFAP) as a Biomarker of Acute Focal Brain Injury

**DOI:** 10.1101/2024.10.11.24315320

**Authors:** Nil Saez-Calveras, Alexander Asturias, James Yu, Barbara Stopschinski, Jaime Vaquer-Alicea, Padraig O’Suilleabhain, Marc I. Diamond, Bhavya R. Shah

## Abstract

**Objective:** The validation of acute brain injury biomarkers has encountered challenges such as the absence of pre-insult measurements, variability in injury timing and location, and interindividual differences. In this study, we addressed these limitations by using Magnetic Resonance-guided High-Intensity Focused Ultrasound (MRgHIFU) thalamotomy to assess plasma biomarker changes after an acute focal brain injury.

**Methods:** This prospective study included 30 essential tremor (ET) and tremor-dominant Parkinson’s disease (TDPD) patients undergoing MRgHIFU thalamotomy at a single academic institution. Blood samples were collected at three specific time points: pre-procedure, 1-hour post-procedure, and 48 hours post-procedure. Plasma levels of glial fibrillary acidic protein (GFAP), neurofilament light chain (NfL), amyloid-beta (Aβ40 and Aβ42), and phosphorylated tau 181 (pTau-181) were measured using the Quanterix Single Molecule Arrays (SiMoA) assay.

**Results:** GFAP levels significantly increased 48 hours post-MRgHIFU in all patients with a thalamotomy lesion. GFAP levels were highly sensitive (89.7%) and specific (96.6%) in detecting the presence of a HIFU lesion with a cutoff value of 216.2 pg/ml. NfL, Aβ40, and Aβ42, also showed statistically significant increases post-procedure but were less robust than GFAP. No changes were observed in pTau-181 levels post-MRgHIFU.

**Conclusion:** Plasma GFAP emerged as a highly sensitive and reliable biomarker for detecting acute brain injury following MRgHIFU thalamotomy. The significant post-procedure elevation of GFAP suggests its potential as an early diagnostic tool for focal brain injuries, particularly in acute stroke. Further research is needed to validate the GFAP injury cutoff identified in this study and to explore its broader clinical utility in the early detection of focal brain lesions.

## Introduction

Plasma biomarkers of neurological injury offer significant advantages in clinical practice, including low cost, rapid turnaround time, and broad accessibility. Together these factors can substantially impact time to treatment and clinical decision making^1^. This is particularly relevant in the context of acute stroke, where timely and accurate diagnosis is critical. Currently, no blood-based biomarkers for acute stroke are available in clinical practice^2^. The development of reliale stroke biomarkers could therefore facilitate timely diagnosis and guide acute treatment interventions such as thrombolytic therapy, especially in cases where routine emergent MR imaging is not possible.

Circulating levels of glial fibrillary acidic protein (GFAP)^3^, and neurofilament light chain (NfL)^4^, have emerged as promising biomarkers of acute and subacute brain injury^5,6^. GFAP is an intermediate filament-III protein primarily expressed in astrocytes which serves as a marker of astroglial activation and injury^7^. Meanwhile, NfL is a neuron-specific cytoskeletal component that reflects axonal damage^8^. Circulating amyloid-β peptides, which are cleavage products of the amyloid precursor protein, and phosphorylated tau species, which are modified forms of the microtubule-associated protein tau, have been proposed as markers of neurodegeneration.^9,10,11,12^. However, they can also increase in acute stroke^13^. Identifying the most reliable and consistent biomarkers of brain injury in acute stroke faces a multitude of challenges. These include the lack of pre-insult biomarker measurements^6,14^, difficulties in accurately determining the timing of injury^15^, and the variability arising from interindividual differences such as co-morbidities, and peripheral tissue sources of these biomarkers^16,17^. Addressing these challenges is essential to validate these plasma biomarkers for use in clinical practice.

Magnetic resonance-guided high-intensity focused ultrasound (MRgHIFU) thalamotomy represents a unique opportunity in addressing these limitations. As an FDA-approved, incisionless therapy for essential tremor (ET)^18^ and Parkinson’s disease (PD), this treatment ablates the dentatorubrothalamic tract (DRTT)/ventral intermediate nucleus (VIM) of the thalamus providing real-time tremor relief.^19^ MRgHIFU ablation also results in temporary disruption of the blood brain-barrier (BBB)^20,21^. We have an established MRgHIFU program, and ET and PD patients routinely undergo MRgHIFU ablation using a precision-imaging based approach named four tract tractography^22,23^. This approach improves clinical outcomes and reduces adverse effects by using patient-specific tractography instead of stereotactic coordinates^23^. At the same time, MRgHIFU with four tract tracrography provides a precise and consistent ablation of a circumscribed anatomic area in the DRTT/VIM of the thalamus across different patients, with minimal target heterogeneity^23^.

By utilizing the controlled timeline and spatial precision of MRgHIFU thalamotomy, we can closely examine the relationship between specific plasma biomarkers and a brain injury that closely resembles that seen in subcortical stroke. This approach eliminates the biases introduced by time variability and interindividual differences that complicate other biomarker studies. The ability to measure biomarkers before, immediately after, and several hours post-injury offers valuable insights into how these biomarkers change over time, providing a clearer understanding of the injury response and pinpointing the source of biomarker release. Prior studies in murine models have explored the changes in NfL, phosphorylated tau 181 (p-tau-181) and other biomarkers associated with BBB opening using focused ultrasound^24,25^. However, no study has evaluated their trajectories in humans after MRgHIFU thalamotomy.

In this study, we compared changes in circulating levels of GFAP, NfL, amyloid-beta species (Aβ40, Aβ42), and p-tau-181 at three specific time points: before, 1 hour after, and 48 hours after MRgHIFU thalamotomy. To our knowledge, this is the first study to assess plasma biomarker changes in human subjects following MRgHIFU ablation. Our findings suggest that MRgHIFU thalamotomy leads to systemic biomarker changes across individuals and represents a powerful tool for acute brain injury biomarker discovery.

## Materials and methods

### Patient information

All patients voluntarily consented to participate in this study. We studied 30 patients who underwent MRgHIFU from January 2023 through March 2024 at a single academic institution (University of Texas Southwestern Medical Center, UTSW). Consent was obtained according to the Declaration of Helsinki^26^ and it was approved by the ethical committee at UTSW. Patients were consented to provide their demographic information, diagnosis, MRgHIFU treatment details, and to undergo intravenous blood collection before and after the procedure. The patient data was collected in a prospective manner and included their basic demographics (gender, age, and dexterity), disease diagnosis, duration and features of the tremor, and prior medication history. The MRgHIFU procedure details were also collected, and included the treatment site, skull density ratio (SDR), number of sonications, mean and maximum temperature (T_max,_ °C) reached, power (W) and energy (J) delivered, history of prior MRgHIFU, treatment response at 48 h, procedure side effects, and MR imaging findings post-procedure.

### Blood processing

Blood collection was performed with purple-top K2 EDTA coated tubes immediately prior (baseline) to MRgHIFU and 1 hour after MRgHIFU in the FUS suite at UTSW Medical Center. The 48-hour time point was collected at a follow-up outpatient appointment. The samples were stored on ice for transport until further processing. The tubes were then spun down at 3900 rpm for 10 minutes at 4 °C in a Beckman Coulter Allegra V-15R centrifuge. The plasma supernatant was then collected and aliquoted into O-ring screw cap 1 ml sterile tubes and stored in a −80 °C freezer until further processing.

### Biomarker detection using SiMoA

The samples were then transported to the UTSW Microarray Core for neurological biomarker detection via the Quanterix Single Molecule Arrays (SiMoA) assay (Billerica, MA). The Quanterix Neurology 4-plex E (NfL, GFAP, Aβ40, Aβ42), and p-tau-181 assays were used in this study. These assays employ a bead-based enzyme-linked immunosorbent assay (ELISA) technology, on which the immunocomplexes formed on single beads containing primary antibody and detection antibody, are isolated in arrays of 50-femtolitere reaction chambers, allowing for the detection of single protein molecules by fluorescence imaging ^27^.

### MRI acquisition for MRgHIFU

For all patients, a 60-minute MRI scan with diffusion tensor imaging (DTI) was performed on a Phillips 3 T MR Scanner (Philips, Best, The Netherlands). The sequences include isotropic T2-weighted (T2W) three-dimensional turbo field echo [field of view (FOV) 24 cm, matrix 268 × 268 mm, repetition time (TR) = 2500, echo time (TE) = 255.56, thickness 0.9 mm, gap = 0 mm, spacing = 0.9 mm], fast grey matter acquisition T1 inversion recovery (FGATIR) (FOV 25 cm, matrix 256 × 256, TR = 6.615, TE = 2.949, thickness = 0.9 mm, gap = 0, spacing = 0.9 mm), axial three-dimensional T1 turbo field echo (FOV = 24 cm, matrix 268 × 187, TR = 8588 ms, TE = 3.93 ms, gap = 0, spacing = 0.9 mm) and 32 direction DTI (FOV 24 × 24 × 15 cm, matrix 96 × 96, B value = 800, TR = 3400, TE = 84.5, acquisition voxel = 2.5 mm, thickness 2.5 mm, spacing 2.5 mm, gap = 0, SNR = 0.99, slices = 60 and Halfscan factor = 0.84).

An identical post-procedure MR imaging was completed on a Phillips 3 T MR Scanner to evaluate the MRgHIFU ablation lesion.

### Statistical analysis

Statistical analyses were completed using GraphPad Prism. One-way analysis of variance (ANOVA) was used for comparison of biomarker levels across the different time points. Tukey’s multiple comparisons test was used for head-to-head comparisons between each time point.

### Data availability

The authors confirm that the data supporting the findings of this study are available within the article and its supplementary material.

## Results

### Patient characteristics

30 subjects were included in this study. The average patient age was 72.1 years (SD 8.6). 22 males (73.3%) and 8 females (26.7%) underwent MRgHIFU thalamotomy. 8 patients (26.7%) received treatment with right-sided DRTT/VIM ablation, and 22 (73.3%) underwent left-sided ablation. 25 of the treated patients had a principal diagnosis of essential tremor (ET), 2 had a diagnosis of ET + tremor-dominant PD (TDPD), and 2 were diagnosed with TDPD. 1 of the subjects had a concurrent diagnosis of right handwriting tremor and dystonia. 5 of the patients had undergone a prior thalamotomy procedure on the contralateral side, and one had received prior treatment on the ipsilateral side with suboptimal response requiring re-treatment. All the treated patients had successful ablations without incident except one of them (*Case #9*), who was unable to complete the MRgHIFU ablation due to skull density considerations and nausea/vomiting during the procedure. In this patient the treatment was terminated, and the post-operative MRI demonstrated a lack of an ablative lesion. Patient characteristics and the MRgHIFU treatment details are summarized in **Table 1** and **Supplementary Table 1**, respectively.

**Table 1:** Summary of patient demographics, tremor diagnosis and characteristics.

|  |  |  |  |  |  |  |
| --- | --- | --- | --- | --- | --- | --- |
| ID# | Gender | Dexterity | Diagnosis | Duration | Characteristics | Prior medication |
| 1 | Male | R | ET | 20+ y. | Bilateral | Primidone, topiramate, propranolol |
| 2 | Male | R | ET | 15 y. | Bilateral. L>R | Propranolol, primidone |
| 3 | Male | L | ET | 35 y. | Bilateral. L>R | Primidone, gabapentin, propranolol |
| 4 | Male | L | ET | 20+ y. | Bilateral | Propranolol, primidone, topiramate |
| 5 | Male | R | ET | 20+ y. | Bilateral. L>R | Propranolol, primidone, topiramate |
| 6 | Male | R | ET | 30+ y. | Bilateral. R>L | Primidone, gabapentin, topiramate |
| 7 | Male | R | Writing tremor + dystonia | 10+ y. | R writing tremor | Primidone, propranolol |
| 8 | Male | R | ET + dystonic head tremor | 30 y. | Bilateral. L>R | Gabapentin, primidone, botulinum toxin (dystonia), clonazepam, propranolol, topiramate |
| 9 | Female | R | ET | 20+ y. | Bilateral. Symmetric | Primidone, propranolol |
| 10 | Male | R | ET, orthostatic syncope | 20+ y. | Bilateral. Symmetric | Propranolol, primidone, Levodopa/carbidopa |
| 11 | Male | L | ET | 50+ y. | Bilateral. L>R | Primidone |
| 12 | Female | R | ET | 10+ y. | Bilateral. L>R | Clonazepam, primidone, propranolol |
| 13 | Male | R | ET | 12 y. | Bilateral. L>R. Mild head tremor | Primidone, propranolol, zonisamide |
| 14 | Female | R | ET | 35 y. | Bilateral. Symmetric. | Propranolol, primidone, topiramate, gabapentin |
| 15 | Male | R | ET | 10 y. | Bilateral. L>R. Mild resting tremor | Primidone, gabapentin, levodopa/carbidopa for resting tremor |
| 16 | Male | R | Severe ET +TDPD | ET: 30+ y.<br>PD: 5 y. | ET: R>L action tremor<br>PD: Resting tremor | ET: Primidone, propranolol<br>PD: Levodopa/carbidopa ER for resting tremor |
| 17 | Female | R | ET | 50 y. | Bilateral + axial + voice tremor. Head, chin, jaw tremor | Propranolol, primidone, clonazepam, topiramate, gabapentin |
| 18 | Male | R | ET | 16 y. | Bilateral. R>L. | Propranolol, primidone, gabapentin, topiramate |
| 19 | Female | R | ET | 30-40 y. | Bilateral. R>L. | Propranolol, gabapentin, *Cala Trio* |
| 20 | Male | R | ET | 30 y. | Bilateral. L>R | Primidone, propranolol |
| 21 | Male | R | ET | 20-30 y. | Bilateral. R>L. | Primidone, gabapentin, propranolol, topiramate, clonazepam |
| 22 | Male | R | ET | 10 y. | Bilateral. | Primidone, topiramate, gabapentin |
| 23 | Female | R | ET | 10+ y. | Bilateral L>R | Zonisamide, primidone |
| 24 | Female | R | TDPD | 20 y. | Resting tremor | Levodopa, trihexyphenidyl |
| 25 | Male | R | ET | 10+ y. | Bilateral. Symmetric | Propranolol, primidone |
| 26 | Male | R | TDPD | 5 y. | Resting tremor | Levodopa, amantadine, trihexyphenidyl |
| 27 | Male | R | ET | 30 y. | Bilateral. Symmetric | Propranolol, topiramate, phenobarbital, hydroxyzine |
| 28 | Male | R | ET + writing tremor | 30 y. | R writing tremor | Propranolol, primidone |
| 29 | Male | R | ET +TDPD | 9 y. | Bilateral action and resting tremor | ET: Primidone<br>PD: Carbidopa/levodopa, rasagiline |
| 30 | Female | L | ET | 14 y. | L>R writing difficulty. Head and jaw tremor | Propranolol, primidone, topiramate, gabapentin |
| **Abbreviations:** ET, Essential Tremor; TDPD, Tremor Dominant Parkinson's disease; L, Left; R, Right; y., years |  |  |  |  |  |  |

### The levels of GFAP, NfL, Aβ40, and Aβ42 increase 48 hours after MRgHIFU delivery

The mean concentration values for plasma GFAP, NfL, Aβ40, Aβ42, and pTau-181 pre-HIFU, 1h post-HIFU and 48h post-HIFU are listed in **Table 2. Supplementary Table 2** includes the biomarker levels for each individual subject. Of note, the 1 h post-HIFU collection for *Case #17* was discarded due to significant hemolysis.

**Table 2:**
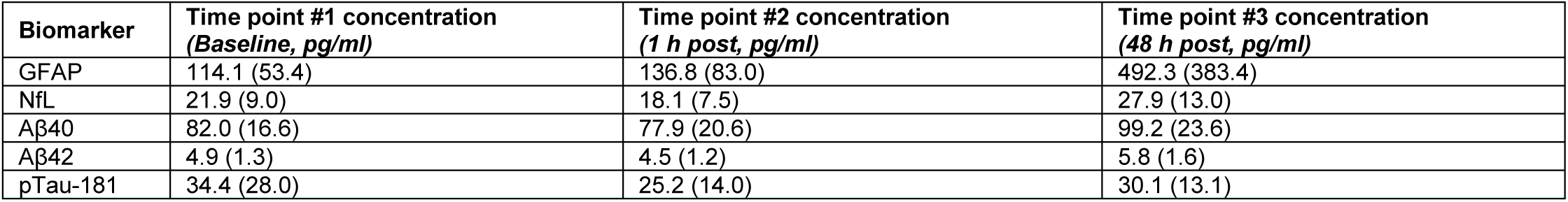
Quantitative measurement of biomarker levels assessed by the Quanterix Single Molecule Arrays (SiMoA) assay.

|  |  |  |  |
| --- | --- | --- | --- |
| Biomarker | Time point #1 concentration<br>(Baseline, pg/ml) | Time point #2 concentration<br>(1 h post, pg/ml) | Time point #3 concentration<br>(48 h post, pg/ml) |
| GFAP | 114.1 (53.4) | 136.8 (83.0) | 492.3 (383.4) |
| NfL | 21.9 (9.0) | 18.1 (7.5) | 27.9 (13.0) |
| Aβ40 | 82.0 (16.6) | 77.9 (20.6) | 99.2 (23.6) |
| Aβ42 | 4.9 (1.3) | 4.5 (1.2) | 5.8 (1.6) |
| pTau-181 | 34.4 (28.0) | 25.2 (14.0) | 30.1 (13.1) |

When the absolute plasma levels of these biomarkers were compared across the three time points (pre, 1h post, and 48h post-HIFU), we observed that plasma GFAP, Aβ40 and Aβ42 levels were significantly higher 48h after MRgHIFU when compared to the baseline and 1-hour post-procedure level. In addition, NfL level was also significantly elevated at 48 hours post-procedure when compared to 1h post (**Figure 1**). No significant differences were observed between the pre- and 1h post-HIFU levels for any of the other biomarkers. Of all the biomarkers, GFAP levels exhibited the largest average increase at 48h post-HIFU [*Pre:* 114.1 pg/ml (SD 53.4) vs. *1h-post:* 136.8 (SD 83.0) vs. *48h-post:* 492.3 (SD 383.4) pg/ml]. Meanwhile, the levels of pTau-181 did not significantly change across any of the time points. In contrast to the rest of cases, *Case #9* did not exhibit a statistically significant change in GFAP levels at 48h. In this patient the MRgHIFU ablation was terminated for treatment intolerance during the procedure. This prevented tissue ablation, which was verified on post-procedure MR (**Figure 2**).

**Figure 1:**
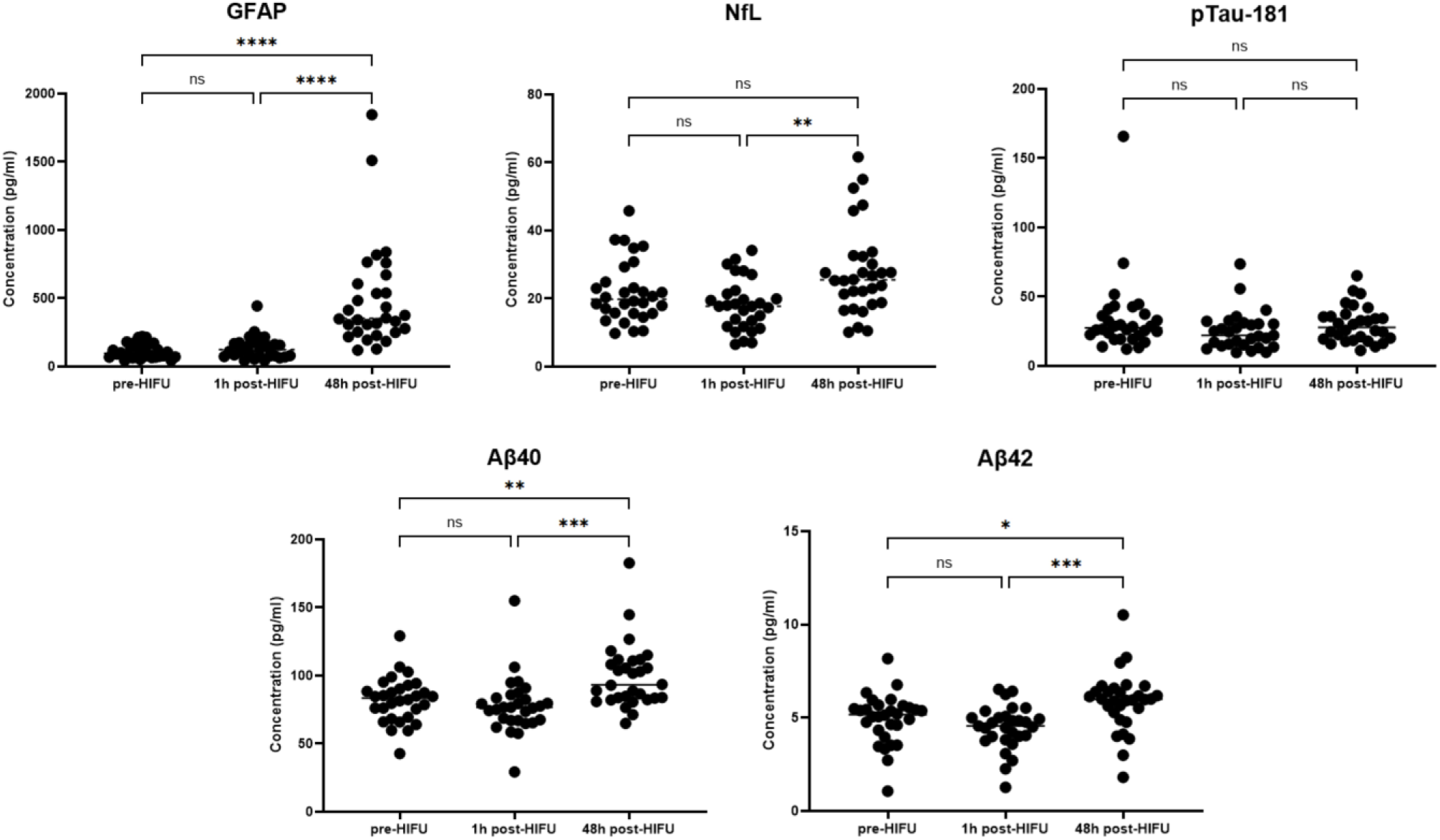
Circulating plasma levels of GFAP, NfL, Aβ40, Aβ42, pTau-181 at baseline, 1 hour post and 48 hours post-HIFU. The levels of GFAP, Aβ40, Aβ42 were significantly elevated after 48 hours when compared to the baseline and 1 hour post-procedure levels. NfL levels were significantly elevated after 48 hours when compared to 1 hour post. No significant change was observed in the levels of p-tau-181. *(*) p-value 0.05; (**) p-value 0.01; (***) p-value 0.001; (****) p-value 0.0001*.

**Figure 2:**
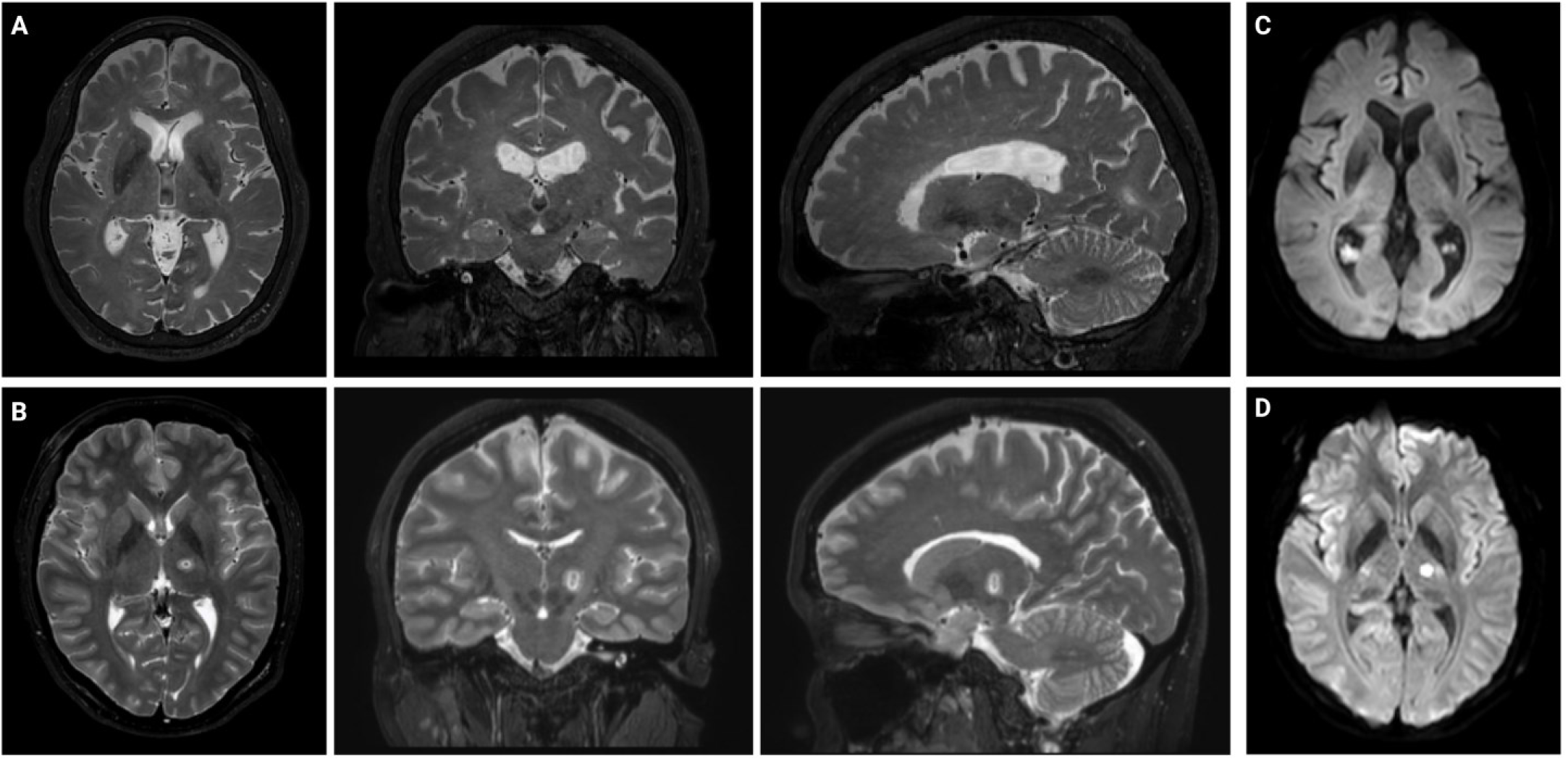
MR imaging results for *Case #9* and *Case #12*. (**A,B**): The axial, coronal and sagittal views of the T2 weighted MR images are shown. No lesion was observed in the post-procedure MR for patient *Case #9* (**A**). A typical MRgHIFU T2 weighted lesion is shown in (**B)** for comparison. **(C,D)**: Axial view of diffusion-weighted imaging (DWI). No diffusion restriction was observed in *Case #9* (**C**), in contrast to *Case #12* (**D**).

**Figure 3** depicts the detailed curves of biomarker trajectories for every individual in absolute values (**A**) As shown, GFAP consistently increased across all the patients who completed MRgHIFU treatment except in *Case #9*. Of note, although the levels of GFAP 1 h post-HIFU did not increase significantly when compared to the pre-HIFU measurement (**Figure 1**), when evaluating the individual GFAP trajectories, a notable increase was observed at 1h post-HIFU in a few subjects. These included *Case #3* (4.19-fold, 444.5 vs. 106.1 pg/ml), *Case #12* (5.76-fold, 255.7 vs. 44.4 pg/ml), *Case #21* (3.45-fold, 217.2 vs. 63.0 pg/ml), *Case #27* (1.85-fold, 127.3 vs. 68.9 pg/ml), *Case #30* (2.14-fold, 157.2 vs. 73.4 pg/ml). No unique characteristics were identified for these patients, except for *Case #3* who had received bilateral thalamotomy.

**Figure 3:**
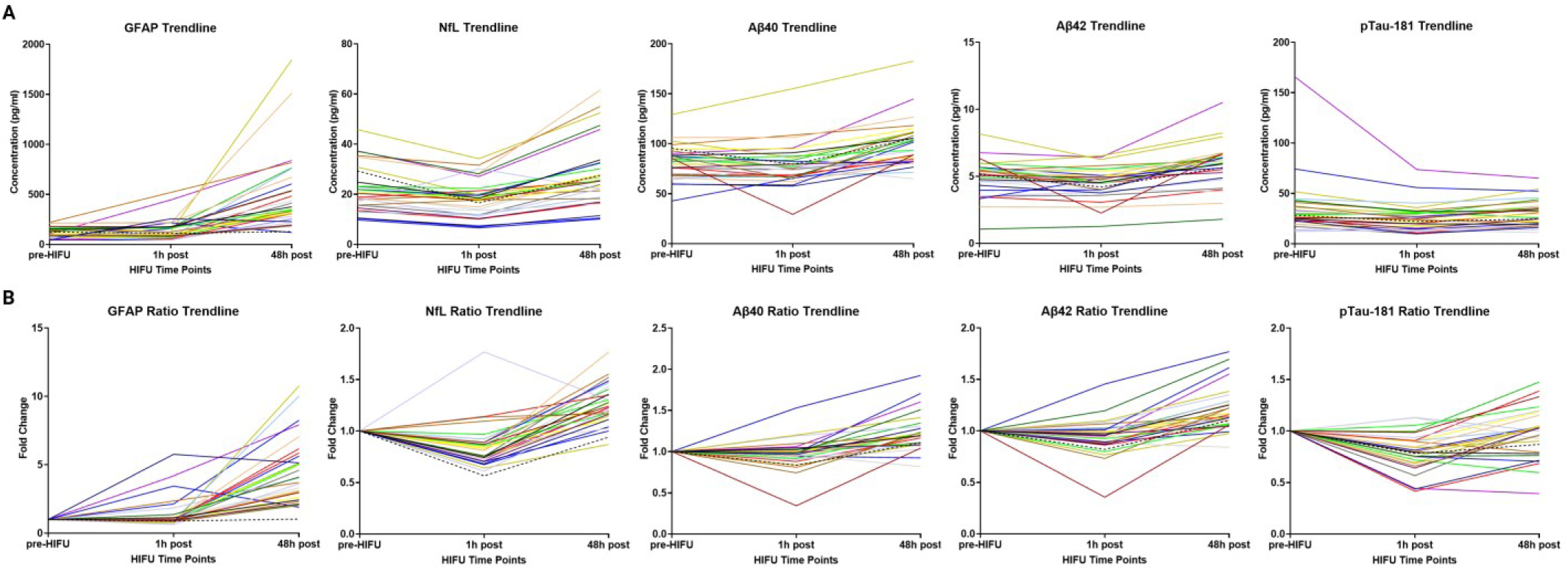
(A). Individual trajectories in the absolute plasma levels of blood biomarkers. (B) Individual trajectories in the fold-change of blood biomarkers. *Case #9*, in whom the procedure was terminated, and no lesion was generated, is identified with a *black dashed line*. GFAP levels were found to consistently increase at the 48-hour time point across all the subjects, except for *Case #9*.

### GFAP serves as a sensitive and specific marker of MRgHIFU ablation after 48 hours

We next tested the sensitivity and specificity of plasma biomarkers to determine the presence of MRgHIFU lesion. *Case #9* was excluded given the absence of a thalamotomy lesion. We conducted a receiver operating characteristic (ROC) analysis by plotting the sensitivity against 1-specificity of our assay for each biomarker, with cases defined as post-HIFU samples (either 48 h post or 1 h post-HIFU), and the controls defined as the baseline pre-HIFU samples (**Figure 4**). GFAP levels at 48h post-HIFU vs. pre-HIFU showed the best ROC fit, with an area under the ROC curve (AUC) of 0.9774 (95% CI 0.945 to 1, p<0.0001) (**Figure 4B**). A GFAP cutoff value of 216.2 pg/ml exhibited a sensitivity of 89.7% (95%CI, 73.6% to 96.4%) and specificity of 96.6% (95%CI, 82.8% to 99.8%) to discriminate the presence of a thalamotomy lesion at 48 h post-HIFU vs. absence of it (pre-HIFU). Using a 224.4 pg/ml cutoff value had a specificity of 100% (95% CI 88.3% to 100%), and a sensitivity of 86.2% (95% CI, 69.4% to 94.5%). Lowering the cutoff to 112.8 ng/ml reached a 100% sensitivity (95% CI, 88.3% to 100%) but with a reduction in specificity to 58.6% (95% CI 40.7% to 74.5%). The sensitivity and specificity for each GFAP cutoff value at the 48 h post-HIFU timepoint vs. pre-HIFU are shown in **Supplementary Table 3.** The rest of biomarkers exhibited a worse ROC fit than GFAP to discriminate presence vs. absence of thalamotomy lesion (**Figure 4A**). However, at the 48h post-HIFU timepoint, the AUC for NfL, Aβ40 and Aβ42 was statistically significant (p value <0.05) (**Figure 4B**). No AUC reached statistical significance at the 1h post-HIFU vs. pre-HIFU timepoint in any of the biomarkers.

**Figure 4:**
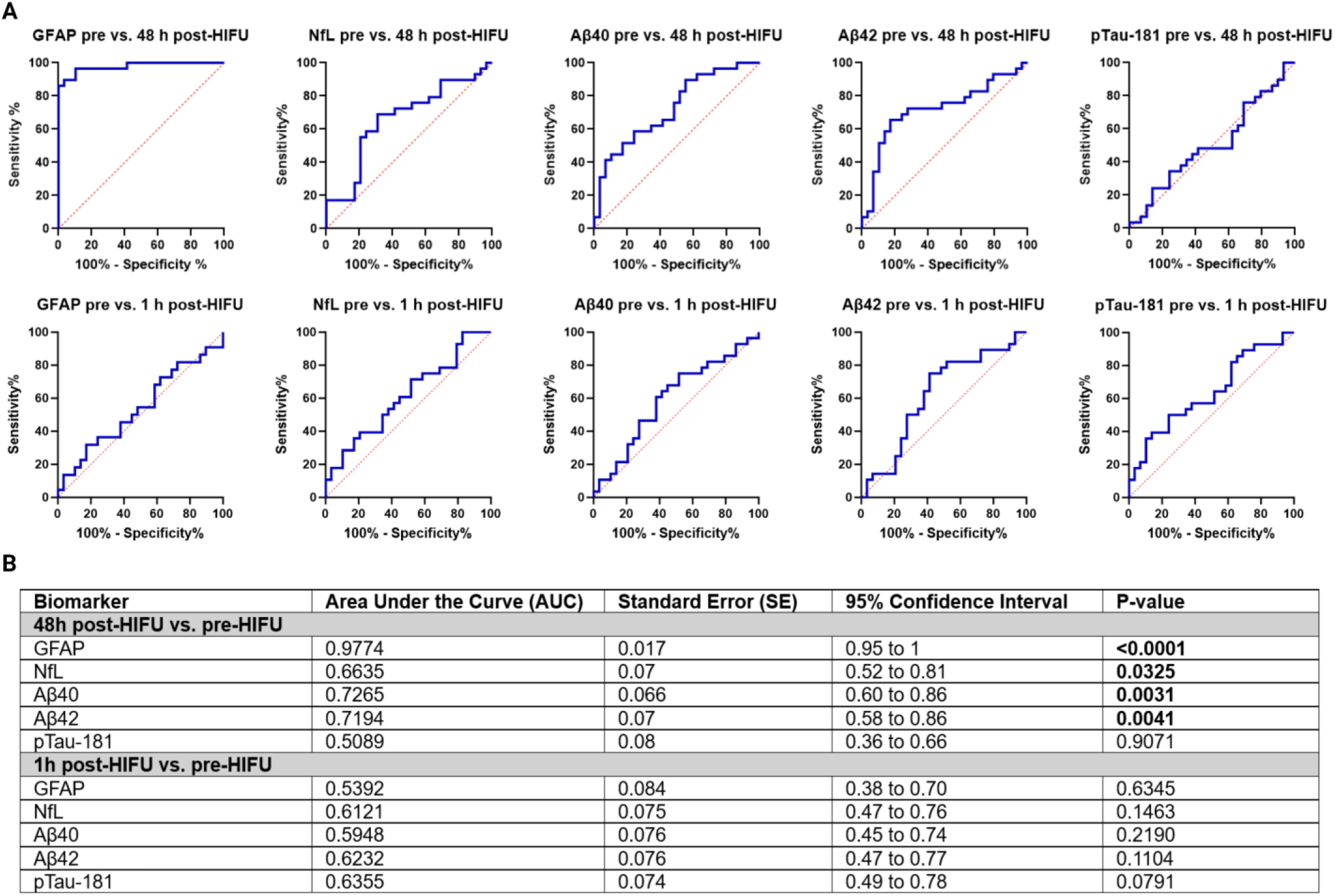
Receiving Operating Curve (ROC) characteristics analysis for each plasma biomarker. **(A).** The results are plotted as 100% - Specificity % vs. Sensitivity % for each biomarker. The levels of biomarkers at 48 h post-HIFU (*top*) or 1 h post-HIFU (*bottom*) were compared against the baseline pre-HIFU level. The GFAP pre vs. 48 h post-HIFU exhibited the best ROC fit. **(B).** Area under the curve, standard error, confidence interval and p-value for all biomarkers at 48h vs. pre-HIFU and 1h vs. pre-HIFU.

### The increase in GFAP levels post-HIFU is higher in second-time MRgHIFU patients at 48 h post-procedure

Next, we tested for differences between those with first-time unilateral MRgHIFU thalamotomy vs. those undergoing a second ablation. 5 subjects had a prior contralateral thalamotomy, and 1 had a prior ipsilateral treatment. The mean difference in treatment time between the first and second procedure was 14.2 months (SD 5.0) with the greatest interval being *Case #18* (22 months), and the shortest being *Case #22* (9 months). *Case #9* was excluded from the analysis in this group, given the suboptimal MRgHIFU treatment.

When the absolute concentrations of biomarkers were measured, baseline and 1 h post-HIFU levels were not significantly different between the two groups, but after 48 hours, there was a significantly higher level of GFAP in those who underwent bilateral thalamotomy (**Figure 5**). However, the interpretation of these findings is limited by the sample size differences between the groups, with 24 patients in the first-time vs. 5 patients in the second-time group.

**Figure 5:**
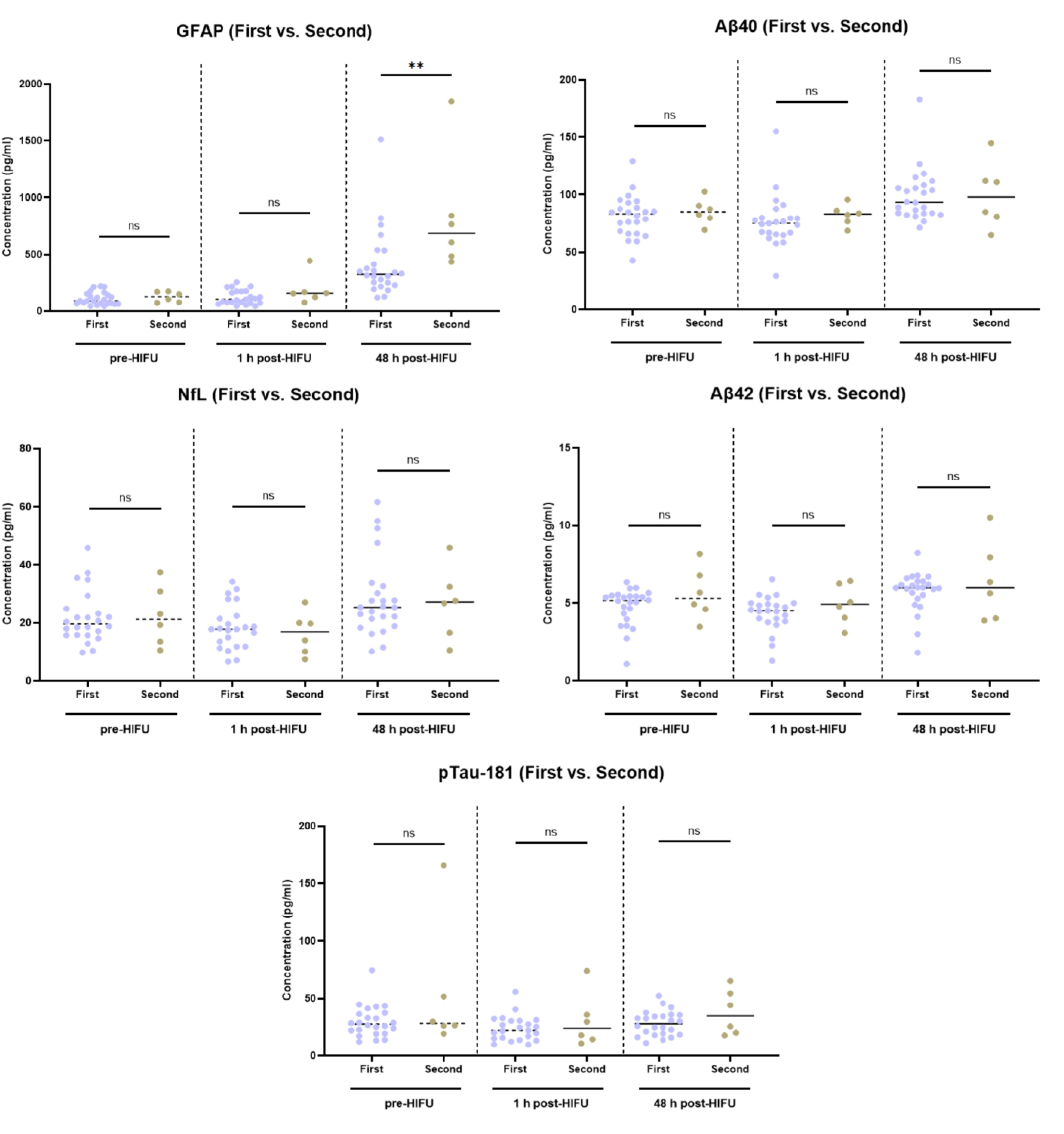
Comparison analysis in absolute biomarker levels between first-time (N=24) and second-time (N=5) thalamotomy patients. Biomarkers GFAP, NfL, Aβ40, Aβ42, p-Tau-181 levels were compared in first-time and second-time thalamotomy patients. Significantly higher levels of GFAP (**) were observed in the second-time thalamotomy patients at 48h post-HIFU. *(*) p-value 0.05; (**) p-value 0.01; (***) p-value 0.001; (****) p-value 0.0001*.

## Discussion

### Plasma GFAP as a consistent biomarker of MRgHIFU lesion

Our study found that MRgHIFU thalamotomy increased the plasma levels GFAP, NfL, Aβ40, and Aβ42 but not pTau-181 at 48 hours after treatment delivery, with GFAP emerging as the most robust biomarker. GFAP levels increased in all individuals who developed a thalamotomy lesion, and notably, there was no increase in GFAP in *Case #9*, where the MRgHIFU delivery was insufficient to generate a lesion. These findings suggest that GFAP is a reliable marker of focal brain injury after MRgHIFU thalamotomy.

Numerous studies have evaluated the use of GFAP as an acute marker of brain injury in acute stroke, as well as in traumatic brain injury (TBI) ^3^. In 2018, the FDA authorized the use of GFAP and ubiquitin carboxy-terminal hydrolase L1 (UCHL1) for clinical use in mild TBI ^3^. In the context of stroke, GFAP has been found to be elevated after both ischemic ^28^ and hemorrhagic injury (ICH) ^29,30^. Some studies have also hypothesized using GFAP to differentiate between ICH and an ischemic injury in the acute setting ^31,32^. In ICH there is a rapid GFAP increase due to the sudden BBB disruption, while in ischemic stroke, a more gradual process is observed due to cytolysis and glial necrosis ^33^. The levels of GFAP in these stroke patients also correlated with stroke severity and a history of a prior stroke ^31^. Other studies have also established GFAP as a possible prognostic marker of acute ischemic stroke. GFAP levels prospectively correlated with clinical and rehabilitation outcomes ^5^. In another study, elevated GFAP on admission after ischemic stroke, predicted poor functional outcomes during the 1-year follow-up ^28^. However, high variation has been observed in all the above studies regarding the GFAP cutoff values used for diagnosis of stroke, which has limited the clinical applicability of this biomarker. Multiple reasons explain this variability, including differences in case vs. control characteristics, timing of injury, and lesion type and location.

### Establishment of a GFAP cutoff value for focal brain injury

One major limitation across all the above studies is the absence of a pre-event measurement that can allow for intraindividual comparison in the change in GFAP and other biomarker levels before and after injury. By using pre- and post-procedural plasma collection in our study we overcame this limitation, which to our knowledge is unprecedented in acute stroke biomarker research. We determine that GFAP levels assessed by SiMoA technology consistently increased across all patients 48 hours after MRgHIFU, and that the use of a GFAP cutoff value of *>216.2 pg/ml* was highly sensitive and specific for detecting the presence of a lesion in these patients. In addition, for a small subset of patients, the levels of GFAP also increased at 1 hour after MRgHIFU. The lesion generated by MRgHIFU highly resembles the features of a subcortical ischemic stroke. The subcortical lesion is also highly consistent between individuals, facilitating the interindividual comparison. Also, GFAP did not elevate in the subject (*Case #9*) in whom the procedure was terminated prior to thermocoagulative necrosis and in whom the neurological effect from the MRgHIFU procedure was transient. This suggests that GFAP may be specific for neuronal death rather than subtotal transient effects as it might be seen clinically in transient ischemic attack (TIA). Our study provides further evidence for the use of GFAP as a marker of CNS subcortical injury and establishes a highly specific and sensitive cutoff value for this biomarker that could be translated to clinical use. Interestingly, we also observed that GFAP levels at 48 hours were higher after second-time vs. first-time thalamotomy. These results align with a prior study which identified higher GFAP levels in acute stroke patients with a history of prior stroke ^31^. However, the interpretation of these results is limited by sample size differences between the groups. Given the reliability of GFAP in detecting small subcortical injuries, it is plausible that GFAP could be even more effective for early detection of cortical strokes, which typically involve larger areas and more extensive tissue damage ^34^. This potential application warrants further investigation and could significantly enhance the clinical utility of GFAP in the context of acute stroke. We suggest that future studies should explore using the established cutoff to determine whether GFAP can serve as a reliable early detection marker in different acute stroke types.

### NfL and Aβ40, Aβ42 also increase after thalamotomy

Plasma NfL, Aβ40 and Aβ42 also increased after 48 hours. However, the changes were not as robust as GFAP and were not present across all subjects, with a significant overlap observed between the three time points (pre, 1h post, 48 h post). The pronounced and consistent elevation of GFAP suggests a rapid and substantial response from astrocytes following the injury, leading to astrocytic activation and reactive gliosis. This early response could be due to the role of astrocytes in maintaining the blood-brain barrier, responding to neuronal injury, and participating in the repair process. In contrast, the more variable increases in NfL, Aβ40, and Aβ42 may reflect ongoing axonal injury or amyloid processing, which may be more dependent on time and lesion size. Plasma NfL quantified within the first 24 hours of stroke has been proposed as a biomarker of stroke outcomes ^35^. However, a systematic review of the temporal trajectory of NfL in stroke determined that the levels of this biomarker significantly increased in the early subacute period after stroke, that is between 14 and 21 days after injury, when compared to the acute setting ^36^. Thus, assessment of time points beyond 48 hours after MRgHIFU could potentially reveal a more consistent elevation of this marker.

### Plasma p-tau-181 does not change after thalamotomy

In contrast to the other biomarkers, we did not observe an increase in pTau-181 following thalamotomy. This lack of a robust pTau-181 response may be attributed to the lesion location, as tau-related pathophysiological changes are less pronounced in the thalamus compared to other regions, and may also take longer to manifest ^37^. While pTau-181 may not be useful as an acute brain injury biomarker, the use of focused ultrasound (FUS) liquid biopsy coupled with pTau species measurement has potential in neurodegenerative conditions. Targeting regions with classic tau pathology, such as the hippocampus, for FUS BBB opening in patients with cognitive impairment may enhance the diagnostic sensitivity of pTau-181 and other neurodegeneration biomarkers, as demonstrated in animal studies ^25^.

### Study limitations

One limitation of our study is the inclusion of only three time points: pre-HIFU, 1 hour post-HIFU, and 48 hours post-HIFU. Notably, the 48-hour time point, where we consistently observed GFAP elevation, is not highly acute. Clinically, in the setting of acute stroke, patients are beyond the window for acute interventions such as thrombolysis at this stage. However, the biomarker’s clinical value remains significant, especially in settings where advanced imaging like MRI is unavailable. In smaller or rural hospitals, where rapid decisions regarding patient transfer for acute stroke care are necessary, a blood test like this could serve as a vital diagnostic tool.

Our study also noted an early GFAP elevation in some patients at 1 hour post-HIFU. However, due to the limited time points used in our study, it is impossible to determine whether a consistent elevation in GFAP occurs earlier than 48 hours across all patients, which could potentially guide acute therapeutic decisions. Therefore, including additional time points between 1 hour and 48 hours, as well as beyond 48 hours, is crucial to gain a more comprehensive understanding of the trajectory and peak levels of biomarkers like GFAP and NfL post-HIFU. This expanded sampling could provide critical insights that inform the development of biomarkers capable of guiding acute interventions in stroke.

## Acknowledgements

We would like to extend our deepest gratitude to the MRgHIFU-treated patients who consented to donate their blood for this study, as well as the medical and nursing staff at the Clements University Hospital Radiology Suite. Without them, this study could not have been possible.

## Supplementary material

‘Supplementary material is available at *Brain* online’.

