## Supplementary Material for "Plasma Glial Fibrillary Acidic Protein (GFAP) as a Biomarker of Acute Focal Brain Injury"

**Table 1S:** Summary of MRgHIFU treatment characteristics

| ID# | **Site** | **SDR** | **N° Sonic** | **T° target reached** | **Mean/ Max T°** | **Power, W** | **Energy, J** | **Prior HIFU?** | **Response at 72h** | **Side effects** | **MR findings** |
|---|---|---|---|---|---|---|---|---|---|---|---|
| 1 | L | 0.61 | 9 | **No** | 56/59 | 1,044 | 25,666 | **No** | Excellent (resolved) | Mild imbalance, lightheadedness | L DRTT ablation. |
| 2 | L | 0.61 | 7 | Yes | 58/61 | 847 | 8,528 | No | Excellent (resolved) | Mild leg imbalance, tongue numbness | L DRTT ablation. Advanced bilateral CVSD |
| 3 | L | 0.60 | 8 | Yes | 55/57 | 1,200 | 24,618 | Yes. R (1 y. 6 mo. prior) | Excellent (resolved) | Imbalance | L DRTT ablation. Prior R DRTT ablation. Periventricular and subcortical CSVD |
| 4 | R | 0.56 | 5 | Yes | 56/58 | 996 | 11,015 | No | Excellent (resolved) | Mild perioral numbness (resolved) | R DRTT ablation. |
| 5 | R | 0.61 | 6 | Yes | 56/57 | 1,095 | 15,415 | No | Excellent (resolved) | Subtle tongue numbness, mild imbalance | R DRTT ablation. Scattered periventricular and subcortical CSVD |
| 6 | L | 0.46 | 6 | Yes | 58/62 | 1,099 | 15,415 | No | Excellent (resolved) | Mild bottom lip numbness | L DRTT ablation. Mild CSVD |
| 7 | L | 0.69 | 6 | Yes | 57/61 | 660 | 7,264 | No | Excellent (resolved) | Mild imbalance | L DRTT ablation |
| 8 | R | 0.69 | 7 | Yes | 57/59 | 896 | 8,987 | Yes. L (10 mo. prior) | Excellent (resolved) | Mild imbalance, mild perioral numbness | R DRTT ablation. Prior L DRTT ablation |
| 9 | L | 0.60 | 3 | **No. Terminated** | 45/46 | 300 | 3,002 | **No** | Tremor 10% improved | **Nausea, vomiting (terminated)** | Small ablation zone in L thalamus with mild restricted effusion. Scattered foci of WM CSVD |
| 10 | L | 0.46 | 4 | **No** | 49/50 | 1,000 | 9,118 | No | Suboptimal response (50% improvement) | None | L DRTT ablation. Mild CSVD |
| 11 | R | 0.55 | 6 | Yes | 55/58 | 798 | 8,773 | No | Excellent (resolved) | None | R DRTT ablation. Mild CSVD, mild diffuse volume loss |
| 12 | L | 0.80 | 7 | Yes | 55/60 | 750 | 8,250 | No | Excellent (95% improved) | Slow walking for balance | L DRTT ablation |
| 13 | R | 0.60 | 6 | Yes | 57/61 | 600 | 6040 | No | Excellent (resolved) | Mild imbalance, improving | R DRTT ablation. Mild volume loss. Scattered CSVD |
| 14 | L | 0.76 | 5 | Yes | 58/62 | 612 | 6080 | No | Excellent (resolved) | Mild-moderate imbalance | L DRTT ablation. Mild volume loss. Mild periventricular and subcortical CSVD |
| 15 | L | 0.62 | 7 | Yes | 56/59 | 850 | 9,413 | No | Excellent (resolved) | Mild imbalance | L DRTT ablation. Scattered subcortical CSVD |
| 16 | L | 0.49 | 4 | Yes | 50/53 | 556 | 7,822 | No | Excellent (95% improved) | R lower lip numbness, mild imbalance | L DRTT ablation. Patchy subcortical CSVD |
| 17 | L | 0.51 | 7 | Yes | 55/56 | 806 | 15,379 | No | Excellent (resolved) | None | L DRTT ablation. Periventricular, WM CSVD. Chronic lacunar infarcts in bilateral corona radiata |
| 18 | R | 0.43 | 7 | Yes | 50/52 | 1,097 | 28,577 | Yes. L (1y. 10 mo. prior) -> 20% return | Excellent (90% improved) | None | R DRTT ablation. Prior L DRTT ablation |
| 19 | L | 0.53 | 7 | Yes | 55/58 | 1,097 | 21,920 | No | Excellent (resolved) | Mild imbalance | L DRTT ablation |
| 20 | L | 0.59 | 6 | Yes | 57/62 | 995 | 11,946 | No | Excellent (95% improved) | Slurred speech, mild R-sided imbalance | L DRTT ablation. Scattered periventricular and subcortical CSVD. Mild global cerebral volume loss |
| 21 | L | 0.75 | 5 | Yes | 56/58 | 671 | 7,020 | No | Excellent (95% improved) | Mild imbalance | L DRTT ablation |
| 22 | L | 0.54 | 6 | Yes | 56/60 | 550 | 9,204 | Yes. R (9 mo. prior) -> 5% tremor return | Excellent (resolved) | Mild bottom lip numbness | L DRTT ablation. Scattered mild CSVD |
| 23 | R | 0.48 | 8 | Yes | 56/59 | 726 | 17,425 | Yes. L (1 y. prior) | Excellent (resolved) | Very mild speech change | R DRTT ablation. Prior L DRTT ablation |
| 24 | L | 0.44 | 9 | Yes | 57/61 | 897 | 12,551 | No | Excellent (95% improved) | Mild-moderate imbalance, speech change | L DRTT ablation. Scattered periventricular, subcortical, splenium of corpus callosum CSVD |
| 25 | L | 0.65 | 7 | Yes | 56/59 | 804 | 11,256 | No | Excellent (95% improved) | None | L DRTT ablation |
| 26 | L | 0.55 | 7 | Yes | 56/61 | 954 | 14,310 | No | Excellent (resolved) | Imbalance, slowed gait | L DRTT ablation |
| 27 | L | 0.57 | 7 | Yes | 54/57 | 799 | 10,393 | No | Excellent (resolved) | Mild imbalance | L DRTT ablation |
| 28 | L | 0.60 | 5 | Yes | 57/60 | 797 | 8,810 | No | Excellent (resolved) | Mild numbness, dysgeusia | L DRTT ablation. Mild volume loss |
| 29 | R | 0.55 | 5 | Yes | 57/60 | 894 | 12,572 | No | Excellent (95% improved) | L perioral numbness | R DRTT ablation. Periventricular CSVD |
| 30 | R | 0.59 | 5 | Yes | 57/60 | 550 | 13,200 | Yes. Right (1 y. 2 mo. prior) -> Suboptimal | Excellent. Mild kinetic tremor, dystonic posture | L perioral numbness | R DRTT ablation. Bilateral CSVD |

**Abbreviations:** CVSD, Cerebral Small Vessel Disease; DRTT, Dentatorubrothalamic tract; h, Hours; J, Joules; L, Left; Max, Maximum; mo., Months; MR, Magnetic Resonance; N° Sonic, Number of Sonications; R, Right; SDR, Skull Density Ratio; T°, Temperature; W, Watts; WM, White Matter

**Table 2S:** Individual patient GFAP, NfL, Aβ40, Aβ42, and pTau-181 biomarker levels at baseline, 1 h post-MRgHIFU, and 48 h post-MRgHIFU

| **ID #** | **GFAP (pg/ml)** | | | **NfL (pg/ml)** | | | **Aβ40 (pg/ml)** | | | **Aβ42 (pg/ml)** | | | **pTau-181 (pg/ml)** | | |
|---|---|---|---|---|---|---|---|---|---|---|---|---|---|---|---|
| | ***Pre*** | ***1h Post*** | ***48 h Post*** | ***Pre*** | ***1h Post*** | ***48 h Post*** | ***Pre*** | ***1h Post*** | ***48 h Post*** | ***Pre*** | ***1h Post*** | ***48 h Post*** | ***Pre*** | ***1h Post*** | ***48 h Post*** |
| 1 | 101.31 | 97.22 | 308.59 | 45.82 | 34.18 | 52.51 | 129.20 | 155.05 | 182.70 | 5.95 | 6.53 | 8.24 | 36.31 | 30.42 | 32.69 |
| 2 | 143.37 | 124.62 | 331.14 | 35.45 | 31.61 | 55.07 | 92.88 | 75.12 | 111.71 | 5.39 | 4.73 | 6.59 | 41.11 | 32.81 | 42.14 |
| 3 | 106.07 | 444.47 | 839.77 | 37.31 | 27.08 | 45.87 | 90.27 | 95.65 | 144.81 | 6.77 | 6.42 | 10.51 | 165.87 | 73.67 | 65.17 |
| 4 | 65.51 | 73.02 | 217.55 | 15.79 | 13.54 | 16.19 | 81.54 | 73.65 | 83.90 | 5.54 | 4.84 | 6.19 | 12.17 | 13.72 | 11.23 |
| 5 | 70.67 | 79.91 | 355.80 | 21.80 | 18.05 | 27.72 | 94.36 | 94.92 | 115.09 | 5.06 | 5.00 | 5.92 | 28.80 | 21.62 | 30.09 |
| 6 | 45.02 | 44.07 | 253.37 | 21.96 | 19.51 | 32.64 | 59.54 | 58.53 | 101.67 | 3.96 | 3.99 | 6.40 | 32.98 | 26.79 | 34.37 |
| 7 | 52.45 | 54.58 | 307.29 | 18.77 | 21.40 | 25.33 | 76.21 | 67.00 | 88.81 | 5.20 | 4.47 | 5.52 | 22.17 | 20.09 | 30.79 |
| 8 | 175.41 | 160.63 | 435.25 | 19.25 | 13.95 | 27.61 | 79.63 | 76.74 | 65.00 | 4.60 | 4.05 | 3.87 | 26.35 | 17.95 | 25.47 |
| 9 | 124.41 | 110.95 | 128.18 | 29.32 | 16.55 | 27.57 | 95.32 | 79.47 | 105.57 | 5.14 | 4.21 | 5.66 | 28.38 | 22.17 | 24.68 |
| 10 | 154.66 | 175.24 | 375.72 | 24.86 | 18.43 | 33.76 | 88.47 | 90.90 | 105.52 | 4.76 | 4.56 | 5.96 | 24.96 | 19.76 | 19.45 |
| 11 | 90.82 | 76.96 | 185.14 | 15.64 | 17.76 | 18.28 | 87.54 | 64.89 | 108.18 | 5.49 | 4.01 | 6.71 | 37.45 | 24.65 | 35.86 |
| 12 | 44.42 | 255.67 | 228.09 | 10.34 | 7.04 | 11.49 | 59.80 | 57.62 | 76.59 | 4.33 | 3.76 | 4.89 | 22.72 | 9.93 | 16.24 |
| 13 | 90.94 | 86.06 | 194.83 | 14.57 | 10.29 | 16.94 | 76.18 | 79.80 | 82.15 | 5.07 | 4.93 | 5.28 | 23.85 | 15.25 | 24.55 |
| 14 | 180.24 | 174.27 | 535.40 | 20.36 | 17.73 | 25.27 | 85.36 | 29.33 | 88.95 | 6.34 | 2.26 | 6.71 | 25.63 | 25.27 | 34.18 |
| 15 | 131.39 | 178.78 | 539.48 | 37.16 | 28.16 | 47.51 | 68.29 | 67.02 | 103.06 | 1.06 | 1.27 | 1.80 | 42.73 | 32.27 | 32.50 |
| 16 | 214.13 | 213.56 | 1511.03 | 34.87 | 28.30 | 61.64 | 106.29 | 106.24 | 126.74 | 2.72 | 2.70 | 2.99 | 32.52 | 27.43 | 37.48 |
| 17 | 220.70 | -* | 818.79 | 17.95 | -* | 21.37 | 99.06 | -* | 118.20 | 5.44 | -* | 6.17 | 26.59 | -* | 21.09 |
| 18 | 150.25 | 168.56 | 765.27 | 23.04 | 19.95 | 32.38 | 82.37 | 83.56 | 110.88 | 4.92 | 4.77 | 6.35 | 29.77 | 29.65 | 43.96 |
| 19 | 166.87 | 163.14 | 349.76 | 21.82 | 18.66 | 25.63 | 84.70 | 77.66 | 103.81 | 5.65 | 4.50 | 6.00 | 28.75 | 30.32 | 35.51 |
| 20 | 69.37 | 43.92 | 250.38 | 18.42 | 11.22 | 22.17 | 84.71 | 76.76 | 92.97 | 4.65 | 4.45 | 6.00 | 13.29 | 12.44 | 15.93 |
| 21 | 62.96 | 217.23 | 119.52 | 9.76 | 6.57 | 10.15 | 42.78 | 65.45 | 82.35 | 3.33 | 4.85 | 5.90 | 74.26 | 55.77 | 52.26 |
| 22 | 171.48 | 124.53 | 1845.50 | 30.82 | 19.72 | 26.77 | 102.66 | 85.89 | 111.85 | 8.17 | 6.26 | 7.95 | 51.67 | 35.71 | 54.31 |
| 23 | 78.46 | 77.60 | 484.48 | 13.50 | 10.12 | 16.54 | 69.44 | 68.72 | 84.87 | 3.47 | 3.07 | 4.01 | 25.94 | 10.81 | 17.77 |
| 24 | 214.86 | 219.02 | 671.58 | 18.48 | 14.98 | 27.64 | 65.93 | 62.10 | 80.99 | 5.36 | 5.00 | 6.77 | 19.37 | 13.24 | 18.47 |
| 25 | 68.47 | 63.02 | 342.23 | 23.15 | 22.39 | 30.18 | 85.03 | 87.52 | 93.59 | 5.98 | 5.53 | 6.40 | 43.32 | 31.14 | 25.74 |
| 26 | 75.83 | 70.55 | 760.14 | 12.82 | 11.83 | 18.82 | 66.09 | 67.61 | 71.35 | 5.37 | 5.54 | 5.98 | 44.60 | 40.37 | 45.64 |
| 27 | 68.88 | 127.27 | 278.88 | 17.06 | 30.17 | 22.17 | 64.06 | 76.22 | 83.54 | 3.53 | 3.82 | 4.76 | 13.95 | 15.79 | 13.94 |
| 28 | 89.95 | 79.59 | 413.02 | 15.67 | 11.78 | 23.86 | 75.53 | 74.37 | 83.85 | 3.52 | 3.59 | 4.11 | 17.22 | 9.76 | 17.94 |
| 29 | 119.80 | 104.74 | 315.90 | 20.69 | 17.34 | 23.00 | 78.68 | 79.35 | 86.54 | 5.44 | 5.36 | 6.14 | 19.11 | 17.16 | 22.72 |
| 30 | 73.41 | 157.16 | 606.06 | 10.52 | 7.39 | 10.52 | 87.46 | 82.38 | 80.73 | 5.68 | 5.07 | 5.63 | 19.32 | 14.48 | 20.14 |

*The 1h post-HIFU sample from Case #17 was discarded due to gross hemolysis

| **Table 3S:** Sensitivity and specificity values for each GFAP cutoff to discriminate between presence (48h post) vs. absence of thalamotomy lesion | | | | |
|---|---|---|---|---|
| **Cutoff Value** | **Sensitivity%** | **95% Confidence Interval** | **Specificity%** | **95% Confidence Interval** |
| > 44.72 | 100 | 88.30% to 100.0% | 3.448 | 0.1769% to 17.18% |
| > 48.74 | 100 | 88.30% to 100.0% | 6.897 | 1.225% to 21.96% |
| > 57.71 | 100 | 88.30% to 100.0% | 10.34 | 3.581% to 26.39% |
| > 64.23 | 100 | 88.30% to 100.0% | 13.79 | 5.497% to 30.56% |
| > 66.99 | 100 | 88.30% to 100.0% | 17.24 | 7.598% to 34.55% |
| > 68.68 | 100 | 88.30% to 100.0% | 20.69 | 9.846% to 38.39% |
| > 69.13 | 100 | 88.30% to 100.0% | 24.14 | 12.22% to 42.11% |
| > 70.02 | 100 | 88.30% to 100.0% | 27.59 | 14.70% to 45.72% |
| > 72.04 | 100 | 88.30% to 100.0% | 31.03 | 17.28% to 49.23% |
| > 74.62 | 100 | 88.30% to 100.0% | 34.48 | 19.94% to 52.65% |
| > 77.15 | 100 | 88.30% to 100.0% | 37.93 | 22.69% to 56.00% |
| > 84.20 | 100 | 88.30% to 100.0% | 41.38 | 25.51% to 59.26% |
| > 90.39 | 100 | 88.30% to 100.0% | 44.83 | 28.41% to 62.45% |
| > 90.88 | 100 | 88.30% to 100.0% | 48.28 | 31.39% to 65.57% |
| > 96.13 | 100 | 88.30% to 100.0% | 51.72 | 34.43% to 68.61% |
| > 103.7 | 100 | 88.30% to 100.0% | 55.17 | 37.55% to 71.59% |
| > 112.8 | 100 | 88.30% to 100.0% | 58.62 | 40.74% to 74.49% |
| > 119.7 | 96.55 | 82.82% to 99.82% | 58.62 | 40.74% to 74.49% |
| > 125.6 | 96.55 | 82.82% to 99.82% | 62.07 | 44.00% to 77.31% |
| > 137.4 | 96.55 | 82.82% to 99.82% | 65.52 | 47.35% to 80.06% |
| > 146.8 | 96.55 | 82.82% to 99.82% | 68.97 | 50.77% to 82.72% |
| > 152.5 | 96.55 | 82.82% to 99.82% | 72.41 | 54.28% to 85.30% |
| > 160.8 | 96.55 | 82.82% to 99.82% | 75.86 | 57.89% to 87.78% |
| > 169.2 | 96.55 | 82.82% to 99.82% | 79.31 | 61.61% to 90.15% |
| > 173.4 | 96.55 | 82.82% to 99.82% | 82.76 | 65.45% to 92.40% |
| > 177.8 | 96.55 | 82.82% to 99.82% | 86.21 | 69.44% to 94.50% |
| > 182.7 | 96.55 | 82.82% to 99.82% | 89.66 | 73.61% to 96.42% |
| > 190.0 | 93.1 | 78.04% to 98.77% | 89.66 | 73.61% to 96.42% |
| > 204.5 | 89.66 | 73.61% to 96.42% | 89.66 | 73.61% to 96.42% |
| > 214.5 | 89.66 | 73.61% to 96.42% | 93.1 | 78.04% to 98.77% |
| > 216.2 | 89.66 | 73.61% to 96.42% | 96.55 | 82.82% to 99.82% |
| > 219.1 | 86.21 | 69.44% to 94.50% | 96.55 | 82.82% to 99.82% |
| > 224.4 | 86.21 | 69.44% to 94.50% | 100 | 88.30% to 100.0% |
| > 239.2 | 82.76 | 65.45% to 92.40% | 100 | 88.30% to 100.0% |
| > 251.9 | 79.31 | 61.61% to 90.15% | 100 | 88.30% to 100.0% |
| > 266.1 | 75.86 | 57.89% to 87.78% | 100 | 88.30% to 100.0% |
| > 293.1 | 72.41 | 54.28% to 85.30% | 100 | 88.30% to 100.0% |
| > 307.9 | 68.97 | 50.77% to 82.72% | 100 | 88.30% to 100.0% |
| > 312.2 | 65.52 | 47.35% to 80.06% | 100 | 88.30% to 100.0% |
| > 323.5 | 62.07 | 44.00% to 77.31% | 100 | 88.30% to 100.0% |
| > 336.7 | 58.62 | 40.74% to 74.49% | 100 | 88.30% to 100.0% |
| > 346.0 | 55.17 | 37.55% to 71.59% | 100 | 88.30% to 100.0% |
| > 352.8 | 51.72 | 34.43% to 68.61% | 100 | 88.30% to 100.0% |
| > 365.8 | 48.28 | 31.39% to 65.57% | 100 | 88.30% to 100.0% |
| > 394.4 | 44.83 | 28.41% to 62.45% | 100 | 88.30% to 100.0% |
| > 424.1 | 41.38 | 25.51% to 59.26% | 100 | 88.30% to 100.0% |
| > 459.9 | 37.93 | 22.69% to 56.00% | 100 | 88.30% to 100.0% |
| > 509.9 | 34.48 | 19.94% to 52.65% | 100 | 88.30% to 100.0% |
| > 537.4 | 31.03 | 17.28% to 49.23% | 100 | 88.30% to 100.0% |
| > 572.8 | 27.59 | 14.70% to 45.72% | 100 | 88.30% to 100.0% |
| > 638.8 | 24.14 | 12.22% to 42.11% | 100 | 88.30% to 100.0% |
| > 715.9 | 20.69 | 9.846% to 38.39% | 100 | 88.30% to 100.0% |
| > 762.7 | 17.24 | 7.598% to 34.55% | 100 | 88.30% to 100.0% |
| > 792.0 | 13.79 | 5.497% to 30.56% | 100 | 88.30% to 100.0% |
| > 829.3 | 10.34 | 3.581% to 26.39% | 100 | 88.30% to 100.0% |
| > 1175 | 6.897 | 1.225% to 21.96% | 100 | 88.30% to 100.0% |
| > 1678 | 3.448 | 0.1769% to 17.18% | 100 | 88.30% to 100.0% |
